# Long COVID is associated with severe cognitive slowing

**DOI:** 10.1101/2023.12.03.23299331

**Authors:** Sijia Zhao, Eva Maria Martin, Philipp A. Reuken, Anna Scholcz, Akke Ganse-Dumrath, Annie Srowig, Isabelle Utech, Valeska Kozik, Monique Radscheidt, Stefan Brodoehl, Andreas Stallmach, Matthias Schwab, Emily Fraser, Kathrin Finke, Masud Husain

**Affiliations:** Department of Experimental Psychology, University of Oxford, Oxford OX2 6GG, UK; Department of Neurology, Jena University Hospital, Jena, Germany; Department of Internal Medicine IV (Gastroenterology, Hepatology and Infectious Diseases), Jena University Hospital, Jena, Germany; Nuffield Department of Clinical Neurosciences, University of Oxford, Oxford OX3 9DU, UK; Center for Sepsis Control and Care (CSCC), Jena University Hospital, Jena, Germany; Department of Psychology, LMU Munich, Munich, Germany; Oxfordshire Post-COVID Assessment Clinic, Oxford University Hospitals Foundation NHS Trust, Oxford, UK

**Author notes:** These authors contributed equally to this work and share last authorship (Kathrin Finke, Masud Husain). **Corresponding Author:** Sijia Zhao Department of Experimental Psychology, University of Oxford, Oxford OX2 6GG, UK.

**Keywords:** COVID-19, post-COVID conditions, Post-Acute COVID Syndrome, cognition, response time, processing speed, attention

## Abstract

**Background:** COVID-19 survivors may suffer from a wide range of chronic cognitive symptoms for months or years as part of post-COVID-19 conditions (PCC). To date, there is no definitive objective cognitive marker for PCC. We hypothesised that a key common deficit in people with PCC might be generalised cognitive slowing.

**Methods:** To examine cognitive slowing, PCC patients completed two short web-based cognitive tasks, Simple Reaction Time (SRT) and Number Vigilance Test (NVT). 270 patients diagnosed with PCC at two different clinics in UK and Germany were compared to two control groups: individuals who contracted COVID-19 before but did not experience PCC after recovery (No-PCC group) and uninfected individuals (No-COVID group).

**Findings:** We identified pronounced cognitive slowing in PCC patients, which distinguished them from age-matched healthy individuals who previously had symptomatic COVID-19 but did not manifest PCC. Cognitive slowing was evident even on a 30-second task measuring simple reaction time (SRT), with PCC patients responding to stimuli ∼3 standard deviations slower than healthy controls. This finding was replicated across two clinic samples in Germany and the UK. Comorbidities such as fatigue, depression, anxiety, sleep disturbance, and post-traumatic stress disorder did not account for the extent of cognitive slowing in PCC patients. Furthermore, cognitive slowing on the SRT was highly correlated with the poor performance of PCC patients on the NVT measure of sustained attention.

**Interpretation:** Together, these results robustly demonstrate pronounced cognitive slowing in people with PCC, which distinguishes them from age-matched healthy individuals who previously had symptomatic COVID-19 but did not manifest PCC. This might be an important factor contributing to some of the cognitive impairments reported in PCC patients.

**Funding:** Wellcome Trust (206330/Z/17/Z), NIHR Oxford Health Biomedical Research Centre, the Thüringer Aufbaubank (2021 FGI 0060), German Forschungsgemeinschaft (DFG, FI 1424/2-1) and the Horizon 2020 Framework Programme of the European Union (ITN SmartAge, H2020-MSCA-ITN-2019-859890).

**Research in context:** *Evidence before this study:* We searched Google Scholar and PubMed for original research or review articles about the cognitive impairment after COVID-19, published up to 3 December 2023. We used terms relating to COVID-19 (SARS-CoV-2, influenza), post-acute symptoms (long COVID, post-COVID conditions, Post-Acute COVID Syndrome) and cognitive impairment (brain fog, cognitive deficit). Previous studies have shown that some people who recovered from the acute symptoms of COVID-19 might nevertheless experience deficits across an array of cognitive functions, including sustained attention, cognitive flexibility, and memory. However, most reports lacked consensus on the precise definition of post-COVID conditions and a common cognitive signature of post-COVID conditions remains unknown.

*Added value of this study:* In this investigation, we identified moderate to severe cognitive slowing in most patients with PCC, but not in most people who previously suffered COVID without developing PCC. This was replicated across two post-COVID clinics in Germany and the UK. To our knowledge, this is the first robust demonstration of cognitive slowing as a cognitive signature of post-COVID conditions.

*Implications of all the available evidence:* Using a 30-second web-based, self-administered psychomotor task, cognitive slowing in PCC can be reliably and easily measured as part of diagnostic work-up, and has potential to be a biomarker to track the progress of rehabilitation of PCC. To encourage researchers and clinicians to employ this task, we have ensured that it is available online with online feedback and all of our code is publicly accessible.

## Introduction

Post-COVID-19 condition (PCC), often known as “long COVID”, is a constellation of chronic symptoms that impair daily functioning and persist for at least two months after the confirmation of SARS-CoV2 infection.^1^ Cognitive symptoms are among the most prevalent features of PCC.^2,3^ People with PCC have now been shown to demonstrate deficits across a wide array of high-level cognitive functions, including sustained attention, cognitive flexibility, and memory.^4^ Such cognitive impairments have been found to correlate with reduced cortical thickness in patients with PCC.^5^

These observations may fit with the general description of their symptoms – "brain fog" – that is commonly used by sufferers, but there is currently a lack of a robust cognitive signature that distinguishes PCC patients from those of other people who suffered SARS-CoV2 infection. This makes it difficult to diagnose with objective markers, and also to develop treatments for cognitive symptoms in this group of patients..

One cognitive abnormality in PCC that has attracted some attention recently is slow processing speed. This has been reported in several investigations conducted in both acute and chronic phases of COVID-19, especially in those with self-reported cognitive symptoms.^6–12^ However, due to the lack of consensus on the precise definition of PCC^13,14^ and vast differences in cognitive task design and administration, it remains unclear whether PCC is associated with generalised cognitive slowing.

Here we aim to test if a very simple, basic deficit – cognitive slowing – is present in people with PCC. We compared PCC patients to healthy people who previously contracted COVID-19 but did not experience PCC, as well as a second control group of healthy people who have never contracted COVID-19 before. In a self-administered psychomotor assessment on their own laptop, PCC patients showed distinctly slower reaction time on a 30-second simple reaction time (SRT) task. Further, performance on this task could predict slowing on a more complex test of sustained attention – the number vigilance test (NVT). Combining the reaction times in these two tasks and questionnaire-derived depression score, predicted with high accuracy whether a previously infected person suffered from PCC.

## Methods

A more comprehensive Methods section can be found in Supplementary Materials.

### Participants

194 patients who fulfilled the National Institute for Health and Care Excellence (NICE) criteria for PCC completed this study. They were diagnosed at the post-COVID center, Department of Internal Medicine and Department of Neurology, Jena University Hospital, Germany (Table 1). For replicating the finding in SRT in the Jena PCC group, we then recruited a second group of patients with PCC (n=76) diagnosed at Long COVID clinic in Oxford, UK. All patients’ SARS-CoV2 infection was confirmed by PCR testing more than 12 weeks ago. Their performance was compared with that of two control groups: The **No-COVID group**, i.e., healthy controls with no COVID-19 history, and the **No-PCC group**, i.e., people who had COVID-19 12 weeks ago but were not experiencing PCC at the time of testing. See Supplementary Materials for more details.

**Table 1.** Participant demographics, questionnaire-derived measures and objective measures of Simple Reaction Task and Number Vigilance Test. . In the Post-COVID condition (PCC) group, all patients met the NICE requirements for PCC. We recruited healthy control participants based on their self-reported health who were healthy and unaware of any neurological conditions. They were then split into two groups based on their responses regarding their history of COVID-19. All participants who self-reported unconfirmed PCC were excluded. All metrics shown in this table are reported as a group mean and 1 standard deviation (SD). T-tests and χ2-tests were used to assess between-group differences, with Bayesian Factor 10 reported as well.<0.001 indicates significant p value which passed multiple-comparison corrections. WHO: World Health Organization. PHQ-9: Patient Health Questionnaire-9. HADS: Hospital Anxiety and Depression Scale. FAS: Fatigue Assessment Scale. BFI: Brief Fatigue Inventory. PSQI: Pittsburgh Sleep Quality Index. ESS: Epworth Sleepiness Scale Sleep Test Questionnaire. PTSD: Post-Traumatic Stress Disorder Test. MWT-B: Multiple Choice Word Test-B for premorbid intelligence. *As control participants claimed to be unaware of any neurological or psychiatric conditions, we may assume that they did not have (pre-)existing conditions.

| Measure |  | PCC | No-PCC | No-COVID | PCC vs No-PCC |  | PCC vs No-COVID |  | No-PCC vs No-COVID |  |
| --- | --- | --- | --- | --- | --- | --- | --- | --- | --- | --- |
|  |  |  |  |  | p value | BF10 | p value | BF10 | p value | BF10 |
| Demographics | n | 194 | 63 | 113 |  |  |  |  |  |  |
|  | Age | 48.8 (10.1) | 48.7 (10.5) | 48.3 (11.5) | 0.92 | 0.16 | 0.64 | 0.14 | 0.8 | 0.18 |
|  | Gender, female, n (%) | 144 (74.2) | 36 (57.1) | 52 (46.0) | 0.01 |  | <0.001 |  | 0.16 |  |
|  | Education, mean years (SD) | 10.6 (1.0) | 14.8 (2.4) | 15.1 (2.5) | < 0.001 | >1000 | < 0.001 | >1000 | 0.41 | 0.25 |
| COVID-19 | WHO COVID-19 Severity Scale | 2.6 (1.0) | 2.1 (0.4) | 0 | < 0.001 | >100 |  |  |  |  |
|  | Time from COVID-19 diagnosis, mean days (SD) | 325.7 (155.2) | 385.6 (199.2) | n/a | 0.014 | 2.67 |  |  |  |  |
|  | Stayed at hospital overnight for COVID-19, yes (% of known n) | 32 (20.4% of 157) | 7 (11.1% of 63) | n/a | 0.1 |  |  |  |  |  |
|  | Pre-existing neurological or psychiatric conditions, yes (% of known n) | 30 (27.3% of 110) | unknown* | unknown* |  |  |  |  |  |  |
|  | Concentration difficulty, yes (% of known n) | 135 (69.6% of 194) | 15 (23.8% of 63) |  |  |  |  |  |  |  |
| Questionnaires | Depression - PHQ-9 | 10.2 (4.8) | 4.7 (5.2) | 4.8 (5.5) | < 0.001 | >1000 | < 0.001 | >1000 | 0.96 | 0.19 |
|  | Depression - HADS | 7.2 (4.7) |  |  |  |  |  |  |  |  |
|  | Anxiety - HADS | 7.4 (4.6) |  |  |  |  |  |  |  |  |
|  | Fatigue - FAS | 33.4 (8.0) |  |  |  |  |  |  |  |  |
|  | Fatigue - BFI | 12.6 (16.9) |  |  |  |  |  |  |  |  |
|  | Sleep quality - PSQI | 8.9 (3.4) | 7.8 (2.8) | 7.7 (2.3) | 0.036 | 1.36 | 0.028 | 1.68 | 0.98 | 0.19 |
|  | Daytime sleepiness - ESS | 13.1 (5.0) |  |  |  |  |  |  |  |  |
|  | PTSD | 36.7 (12.2) |  |  |  |  |  |  |  |  |
|  | Premorbid IQ - MWT-B | 28.9 (4.6) |  |  |  |  |  |  |  |  |
| **Simple Reaction Time** | Mean RT (s) | 0.49 (0.20) | 0.35 (0.10) | 0.33 (0.06) | < 0.001 | >1000 | < 0.001 | >1000 | 0.07 | 0.85 |
|  | Age-adjusted RT (z) | 3.05 (3.50) | 0.59 (2.15) | 0.12 (1.37) | < 0.001 | >100 | < 0.001 | >1000 | 0.12 | 0.56 |
|  | CV of RTs | 0.22 (0.09) | 0.25 (0.20) | 0.22 (0.13) | 0.3 | 0.34 | 0.79 | 0.21 | 0.34 | 0.28 |
|  | Age-adjusted CV (z) | 0.38 (1.13) | 0.56 (2.10) | 0.31 (1.43) | 0.6 | 0.24 | 0.78 | 0.21 | 0.4 | 0.25 |
| **Number Vigilance Test** | Mean RT (s) | 0.61 (0.07) | 0.57 (0.08) | 0.57 (0.07) | < 0.001 | >100 | < 0.001 | >1000 | 0.73 | 0.18 |
|  | Age-adjusted RT (z) | 0.76 (1.16) | 0.09 (1.39) | 0.11 (1.07) | < 0.001 | >100 | < 0.001 | >1000 | 0.94 | 0.17 |
|  | Mean accuracy | 0.70 (0.20) | 0.73 (0.22) | 0.78 (0.16) | 0.34 | 0.24 | < 0.001 | 36.79 | 0.09 | 0.67 |
|  | Normalised accuracy (z) | -0.69 (1.41) | -0.54 (1.75) | -0.08 (1.11) | 0.48 | 0.2 | < 0.001 | >100 | 0.035 | 1.34 |
|  | Normalised change in RT (z) | 0.39 (6.59) | -0.14 (1.76) | 0.04 (1.12) | 0.53 | 0.19 | 0.58 | 0.15 | 0.42 | 0.23 |
|  | Normalised change in accuracy (z) | 0.07 (1.34) | -0.19 (1.28) | -0.09 (1.28) | 0.18 | 0.37 | 0.32 | 0.21 | 0.6 | 0.19 |
|  | On-task tiredness (%) | 56.57 (25.03) | 50.21 (29.59) | 47.33 (26.20) | 0.1 | 0.57 | 0.003 | 10.16 | 0.51 | 0.21 |
|  | Change in tiredness (slope) | 0.18 (1.32) | -0.05 (1.48) | 0.16 (1.19) | 0.24 | 0.3 | 0.92 | 0.13 | 0.29 | 0.29 |

### Ethics

The study was carried out in accordance with the Helsinki II ethics regulations. All participants gave electronic informed consent prior to the experiment. Ethics were approved by the ethics committee of Jena University Hospital (Approval Reference: 5082-02/17) and South Central – Oxford A Research Ethics Committee (Approval Reference: 18/SC/0448).

### Simple Reaction Time Task (SRT)

Participants first completed **SRT**, which required them to press the spacebar when a large red circle appeared in the centre of the screen (**Figure 1A**). The diameter of the circle was scaled as 50% of the screen height. Once participants pressed the spacebar, the red circle disappeared and would reappear after a randomised time interval between 0.5 and 2 seconds. There was a total of 16 trials, with the results of the first two trials omitted from further analysis. The mean and coefficient of variance (CV) were then computed for the remaining 14 trials.

**Figure 1.**
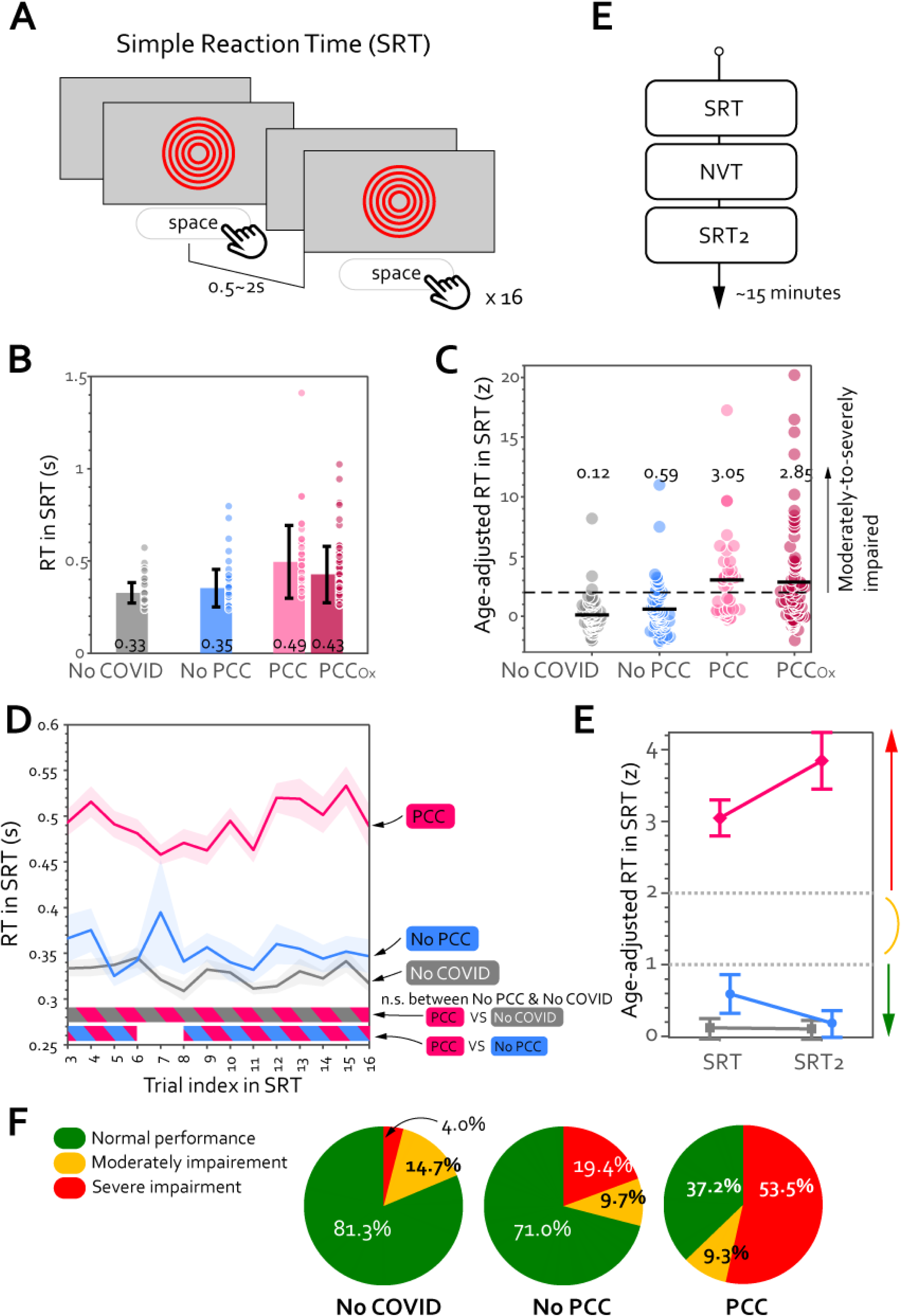
PCC patients were slower than people without PCC, including those who had previously contracted COVID-19. (A) Simple Reaction Time (SRT) task contained a total of 16 trials. The mean RT for each group is shown in (B) with each dot representing individual data and the error bar showing ± 1 SD. Except for the first two trials, which were exceptionally noisy and sluggish, the mean RT was calculated from all trials for each participant. PCC Ox indicates the PCC Oxford group. The value at the bottom of each bar indicates the mean of that group. (C) To account for the effect of age on response speed, all participants’ speed were adjusted based on the No-COVID controls in the same age. Z-score indicates the number of standard deviations from the age-matched normative population. The coloured circles indicate individual results and the black solid line marks the group mean. The horizontal dash black line indicates the threshold for severe impairment (2 SD). (D) Throughout the SRT, PCC participants (pink curve) reacted significantly slower than the other two groups. RT was computed for every trial in SRT across participants and plotted against the trial index. The shaded area shows ± 1 SEM and the horizontal lines at the bottom indicate time intervals where bootstrap statistics confirmed significant differences between two groups (P<0.05). While there was no difference between No-COVID (grey) and No-PCC (blue), PCC (pink) was significantly slower than No-COVID throughout the SRT task (pink-grey stripy horizontal line at bottom) as well as No-PCC (pink-blue stripy horizontal line). (E) To replicate the result seen in SRT, all participants completed the same task again after the second task. (E) The group mean for both sessions of SRT is plotted with group as separate lines. No-COVID (grey line) and No-PCC (blue line) showed a normal performance in both sessions (below the threshold for moderate impairment (> 1 SD). In contrast, PCC (pink line) showed severe impairment (> 2 SD) in both sessions. The error bar indicates 1 standard error of mean. (F) On the individual level, most of No-COVID and No-PCC controls had a normal speed (< 1 SD) while most of PCC patients showed significant impairment.

This task was performed by 119 PCC patients (age 46.6 (SD 12.2, range 19 to 75, 80 females (67.2%)). In addition, 63 No-PCC participants (age 48.7 (10.5), 36 females), and 75 No-COVID participants (age 46.6 (11.9), 29 females) also completed.

### Number Vigilance Test (NVT)

194 participants completed the NVT, an online visual sustained attention task described in the preceding study.^15^ Participants were required to maintain alertness, monitor a rapidly changing stream of numbers, and press the spacebar when they spotted the uncommon target "0". Try it in both English and German at [https://octalportal.com/pcc/]. After every minute, participants reported their level of fatigue (“How tired do you feel now?”) and motivation (“How motivated do you feel?”) using a visual analogue scale. Accuracy was computed as true positive rate. Change of reaction time/accuracy was computed as the slope of the first-degree polynomial curve fitted for the nine minutes (9 time-points), using MATLAB function polyfit.

Both tasks were implemented using PsychoPy v2021.1.2. and hosted on pavlovia.org. All participants used Chrome browser on a desktop/laptop computer with a keyboard.

### Questionnaires

All participants completed two questionnaires for measuring depression level (Patient Health Questionnaire-9, PHQ-9) and their sleep quality (Pittsburgh Sleep Quality Index, PSQI).

All PCC patients also completed:

● Hospital Anxiety and Depression Scale (HADS-D)^16^
● Fatigue Assessment Scale (FAS)^17^
● Brief Fatigue Inventory (BFI)^18^
● Epworth Sleepiness Scale (ESS)^19^
● Post-traumatic stress-scale-14 (PTSS-14)^20^
● Mehrfachwahl-Wortschatz-Intelligenztest (MWT-B) for Premorbid IQ^21^

See more details in Supplementary Materials.

### Statistical analysis

For analysis and data visualisation purposes MATLAB (version R2023a) and R studio (version 12.0) were used.

P values for all group comparisons (t-test for continuous variables or χ2 test for categorical variables) were adjusted with Bonferroni correction. Bayesian factor (BF10) was reported when applicable. A non-parametric bootstrap-based statistical analysis was used to identify time intervals in which groups exhibit differences.^22^ All reported bivariate correlations were performed using Pearson’s correlation method. Kendall’s correlation method is used for correlation between a continuous measure (e.g., reaction time) and an ordinal measure (e.g., WHO COVID-19 severity scale). To investigate the effect of mental health and group on cognitive slowing, the questionnaire-derived mental health metrics were first z-scored across all participants, and generalised linear models (GLM) were conducted in MATLAB, using the function fitglme.

Group classification prediction used MATLAB-based algorithms for the purpose of feature ranking to estimate the absolute contribution of each metric. The fscchi2 function in MATLAB was used to predict group classification and produce the rank’s importance scores. All importance scores were then transformed into p-values by calculating the exponential of their negative value. Group classification was done through multiple logistic regression using MATLAB function fitglm and its ROC curve was plotted using the MATLAB function perfcurve.

## Results

### Severe psychomotor slowing in PCC

Participants first completed **SRT** (**Figure 1A**). Regardless of prior COVID-19, the average reaction time (RT) for healthy controls (collapsed across No COVID and No PCC groups) was 0.34±0.01 seconds. In contrast, PCC patients responded significantly more slowly, with a mean of 0.49 seconds (**Table 1**; **Figure 1B**).

Psychomotor speed prolonged with healthy ageing (RT positively correlated with age amongst controls: Pearson r=0.27, p=0.0014). Therefore, we accounted the effect of age for all objective metrics (e.g., RT in seconds) by computing number of standard deviations from the mean of the No-COVID group in the same age group (± 3 yrs). The mean RT for PCC patients was 3.05±0.02 SDs longer than that of their age-matched No-COVID controls (**Figure 1C**). This significant delay in response was evident from the start of the SRT (**Figure 1D**).

PCC had a normal coefficient of variance in RT (**Table 1**), indicating that their responses were sluggish but not more variable. Individuals with recent (re-)infection were excluded, indicating that this considerable rise in RT cannot be solely attributed to (sub-)acute cognitive alteration following the COVID-19 infection.^4,23–25^

We further replicated this finding with the same individuals 10 minutes later, indexed as SRT2 (**Figure 1D**). PCC patients were substantially impaired compared to the two control groups in both sessions (**Figure 1E**; 2 (session) x 3 (group) ANOVA showed a main effect of group (F(2,354)=49.4, p<0.001, η2=0.22), but no effect of task (F(1,354)=0.13, p=0.7, η2<0.001). Post-hoc comparisons with Bonferroni correction show PCC > No-PCC (difference=3.06±0.37 z, t=8.31, p<0.001) and PCC > No-COVID (difference=3.33±0.36, t=8.2, p<0.001) regardless of the session.

We define any patient with speed slower than 1 SD from the norm average as moderate slowing and those above 2 SD from the average as severe cognitive slowing. In the current sample, 53.5% of PCC patients showed severe cognitive slowing, in contrast to 4.0% in No COVID control group (**Figure 1F**). Overall, PCC group participants had significantly higher proportion of moderate-to-severe impaired cases than No-PCC group (29.1%, χ2(1,N=60)=11.8, p=0.0006) and the No-COVID group (18.7%, χ2(1,N=77)=23.5, p<0.0001).

Out of 194 patients with PCC, 72 completed Montreal Cognitive Assessment (MoCA) in person. They had an average score of 27.7±0.1, ranging between 21 and 30. 12.6% showed poor global cognition, as they scored below the cognitive-impairment cut-off 26. However, there was no correlation between the MoCA score and their cognitive slowing (Pearson’s correlation between MoCA and age-adjusted SRT: r=-0.003, p=0.98; correlation with raw SRT: r=-0.17, p=0.16).

Is this cognitive slowing specific to this group of PCC patients? To answer this question, the same SRT was administered to 76 PCC patients diagnosed in the Long COVID clinic in Oxford, UK (PCC-Ox group, see Supplementary Materials). This cohort also demonstrated significant RT increases (2.83±0.05 z slower than their age-adjusted No-COVID controls) and did not differ from the PCC patients tested in Jena (t(116)=0.29, p= 0.77, BF=0.21, **Figure 1C)**. This replication in a separate centre in a different country provides evidence that the psychomotor slowing is generalised among PCC patients.

### Is cognitive slowing associated with mental health?

Depression and sleep deprivation may increase RT.^26–28^ PCC patients exhibited moderate to severe depressive symptoms and substantially less restful sleep compared with the two control groups (**Table 1**), consistent with previous reports of high prevalence of mood and sleep dysregulation after COVID-19 infection.^29^ However, age-adjusted RT in SRT showed no relationship with self-reported depression (PHQ-9 and HADS), anxiety (HADS), fatigue (FAS and BFI), sleep disturbance (PSQI and ESS), or post-traumatic stress disorder (PTSS-14) (all r values<0.27, p values >0.087). The relationships between cognitive slowing and all questionnaire-derived mental phenotypes are visualised as a network plot in **Figure 2**, in which the strength of the relationship is represented by the distance between the metrics. RT, located at the top of **Figure 2**, showed a clear dissociation from the mental health phenotypes which were closely related to each other, clustering on the right side of the figure.

**Figure 2.**
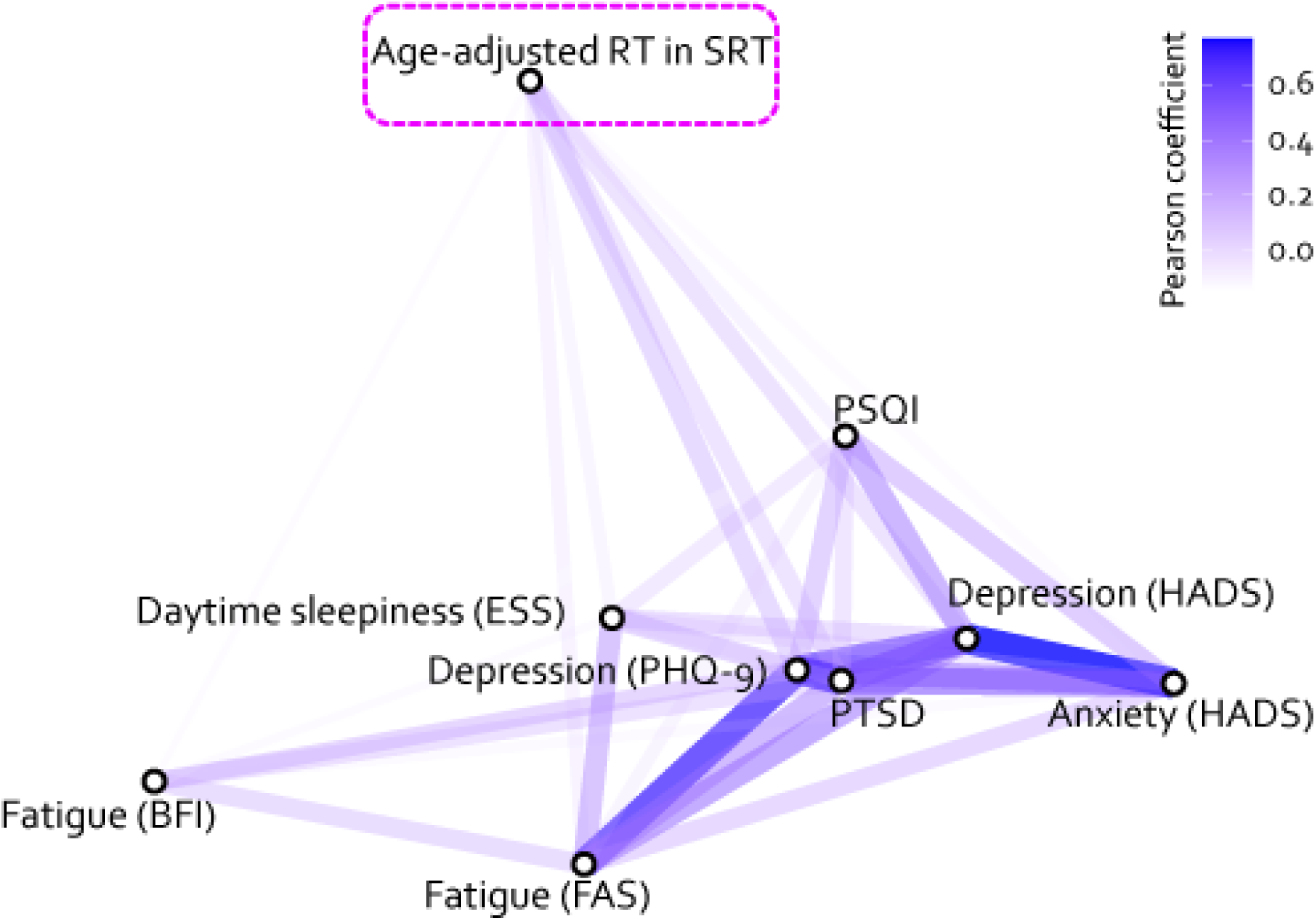
Network plot of relationships between the cognitive slowing (age-adjusted RT in SRT, highlighted with pink dashed rectangle) and all self-reported metrics (bottom cluster) amongst the PCC patients. All depicted relationships are associated with a positive correlation and are rendered in blue. The shorter the distance between two metrics, the stronger their relationship (the higher the correlation coefficient). PHQ-9: Patient Health Questionnaire-9. HADS: Hospital Anxiety and Depression Scale. FAS: Fatigue Assessment Scale. BFI: Brief Fatigue Inventory. PSQI: Pittsburgh Sleep Quality Index. ESS: Epworth Sleepiness Scale Sleep Test Questionnaire. PTSD: Post-Traumatic Stress Disorder Test. Although depression did not predict the cognitive slowing, the combination of depression level and age-adjusted speed in SRT and number vigilance test (NVT) predicts PCC accurately.

This dissociation between cognitive slowing in PCC and mental health symptoms is further supported by a GLM analysis. The GLM examined the relationship of cognitive slowing to the level of depression and the diagnosis (PCC or not) amongst all participants (age-adjusted RT∼PHQ-9*group). The results of this analysis revealed a significant main effect of group [F(1,157)=18.26, p<0.0001], but no interaction between group and depression [F(1,157)=1.83, p=0.18] and no effect of depression [F(1,157)=0.25, p=0.62]. Similarly, no relationship was found with sleep disturbance level (age-adjusted RT ∼ PSQI * group; no interaction [F(1,160)=0.47, p=0.49], no main effect of PSQI [F(1,160)=0.34, p=0.56] but significant effect of group [F(1,160)=29.49, p<0.0001]).

### PCC patients were slow and less vigilant

Next, we asked if the PCC slowing is also present in a cognitively more demanding task than the SRT. To investigate this, we used NVT, a paced vigilance test which emphasises that participants should try to be accurate in their responses, with RT being an implicit measure (**Figure 3A**). Similar to their slowness on the SRT, PCC patients took substantially longer to react to targets compared to healthy controls (**Figure 3B**), regardless of whether the controls had COVID-19 previously or not (**Table 1**). The slowness was maintained throughout the entire course of nine minutes (**Figure 3C**). Importantly, RT on the NVT was strongly associated with the slowness observed in SRT, as age-adjusted RT in SRT alone could explain 35.4% of variance in age-adjusted RT in NVT amongst PCC patients (F(1,179)=30.7, p<0.001). Overall, PCC patients showed a mild, yet significant, cognitive slowing (**Figure 3D**) and a smaller proportion of PCC patients showed severe impairments in this task (i.e., people whose speed was slower than 3 SD from the healthy controls in their age, **Figure 3E**).

**Figure 3.**
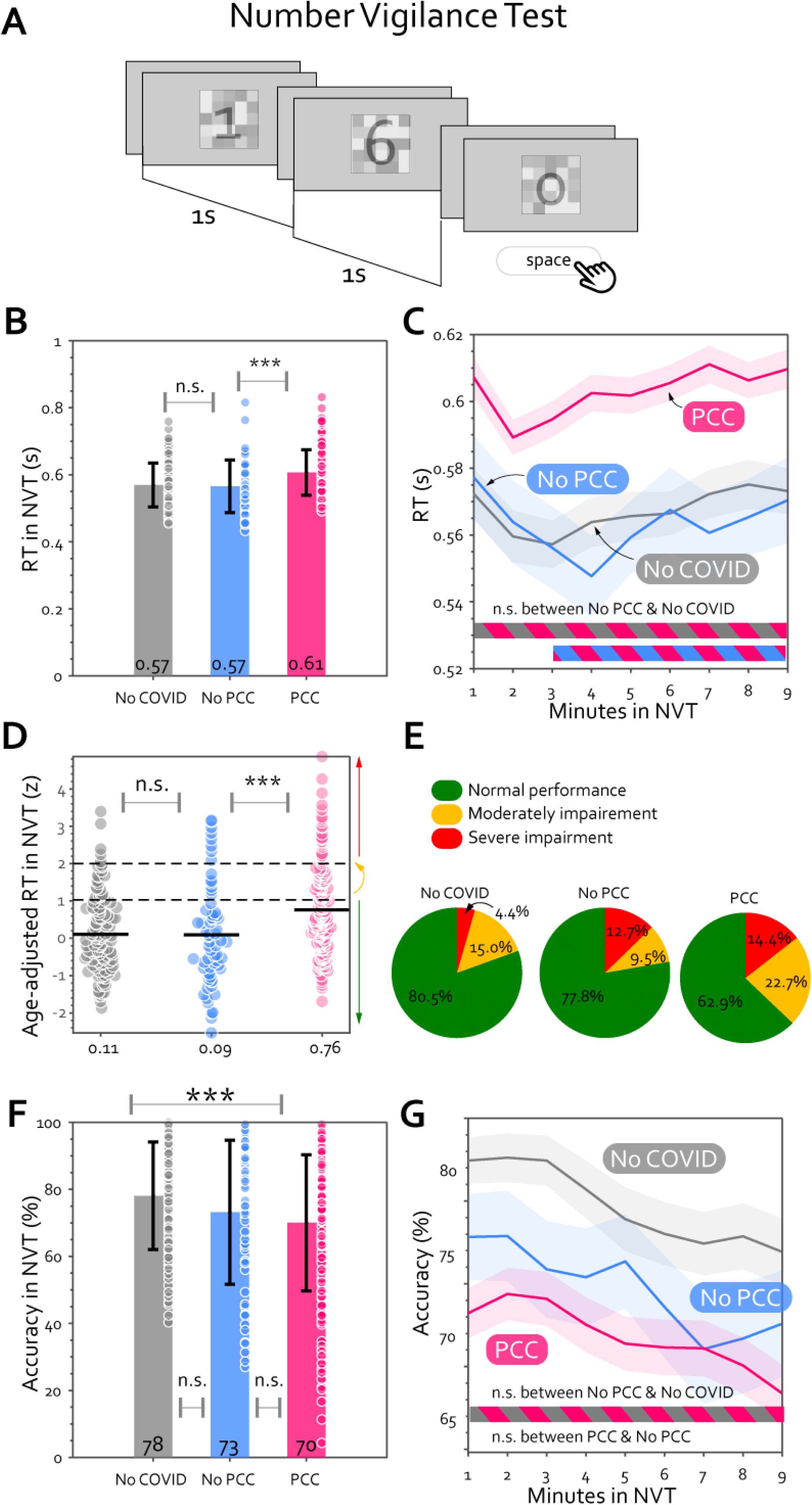
PCC patients responded slower and worse at detecting targets in the Number Vigilance Test (NVT). (A) Following the SRT, all participants completed the NVT, in which a number between 0 and 9 was displayed at 1 Hz and it was displayed for 0.1 seconds while being masked by a transparent checkerboard. Participants were instructed not to press anything if the number was between 1 to 9 and press spacebar as soon as possible when seeing the target number 0. The frequency of the target “0” was low (25%) and would not happen consecutively. The mean RT over 9 minutes for each group is shown in (B), and the mean RT for every minute of this test was plotted against the time (C). n.s. means no significant difference. * means p<0.05, *** means p<0.001 and passes multiple comparison corrections. The shaded area shows ± 1 SEM and the horizontal lines at the bottom indicate time intervals where bootstrap statistics confirmed significant differences between two groups (P<0.05). (D) To account for the effect of age on RT in the NVT, all participants’ speed were age-adjusted based on the No-COVID controls in the same age. Z-score indicates the number of standard deviations from the age-matched normative population. The coloured circles indicate individual results and the black solid line marks the group mean. The two horizontal dash black lines indicate the thresholds for moderate (>1 SD) and severe impairments (>2 SD). (E) The pie charts show the proportion of individuals who had a normal speed (< 1 SD, green area), moderate impairment (> 1 SD, yellow area) and severe impairment (> 2 SD, red area) in this test. The mean accuracy during this task is plotted in (F) and against time in (G). PCC (pink) was significantly less vigilant than No-COVID throughout the SRT task (pink-grey stripy horizontal line at bottom).

Again, cognitive slowing on this task could not be explained by depression (GLM of age-adjusted RT with normalised PHQ-9 score and group; no interaction [F(1,271)=0.24, p=0.63], no effect of PHQ-9 [F(1,271)=0.0.04, p=0.84], but significant effect of group [F(1,271)=18.21, p<0.0001]) or sleep disturbance (GLM with normalised PSQI score and group; no interaction [F(1,202)=0.55, p=0.46], no effect of PSQI [F(1,202)=0.06, p=0.80], but significant effect of group [F(1,202)=39.45, p<0.0001].

In addition to slowing, PCC patients were also less vigilant to visual stimuli compared to the uninfected participants (**Figure 3D**, t(305)=-3.95, p<0.0001, BF10=194.7). However, the vigilance level could not distinguish if an infected individual was experiencing PCC or not (no significant difference between PCC and No-PCC groups: t(255)=-0.95, p=0.34 BF10=0.24; **Figure 3F**). PCC patients also showed good maintenance of vigilance over time (**Figure 3G**). Although their accuracy significantly declined over time (t(193)=-3.5, p<0.001, BF10=25.3), the decrement was minimal in magnitude (slope of accuracy over time=-0.005±0.002 minute^-1^), statistically similar to the No-PCC and No-COVID controls (**Table 1**).

Could PCC patients deliberately slow down to maintain accuracy (aka, speed-accuracy trade-off, SAT)? The results showed the converse: RT negatively correlated with accuracy (**Figure 4A**, r≤-0.54, p<0.001 in all three groups), indicating that slower individuals actually had lower vigilance too. This pattern remained true within individuals: PCC patients with a higher tendency for SAT (i.e., blocks with longer RT associated with higher accuracy, **Figure 4B**) were poorer performers overall in both accuracy and speed (**Figure 4C**).

**Figure 4.**
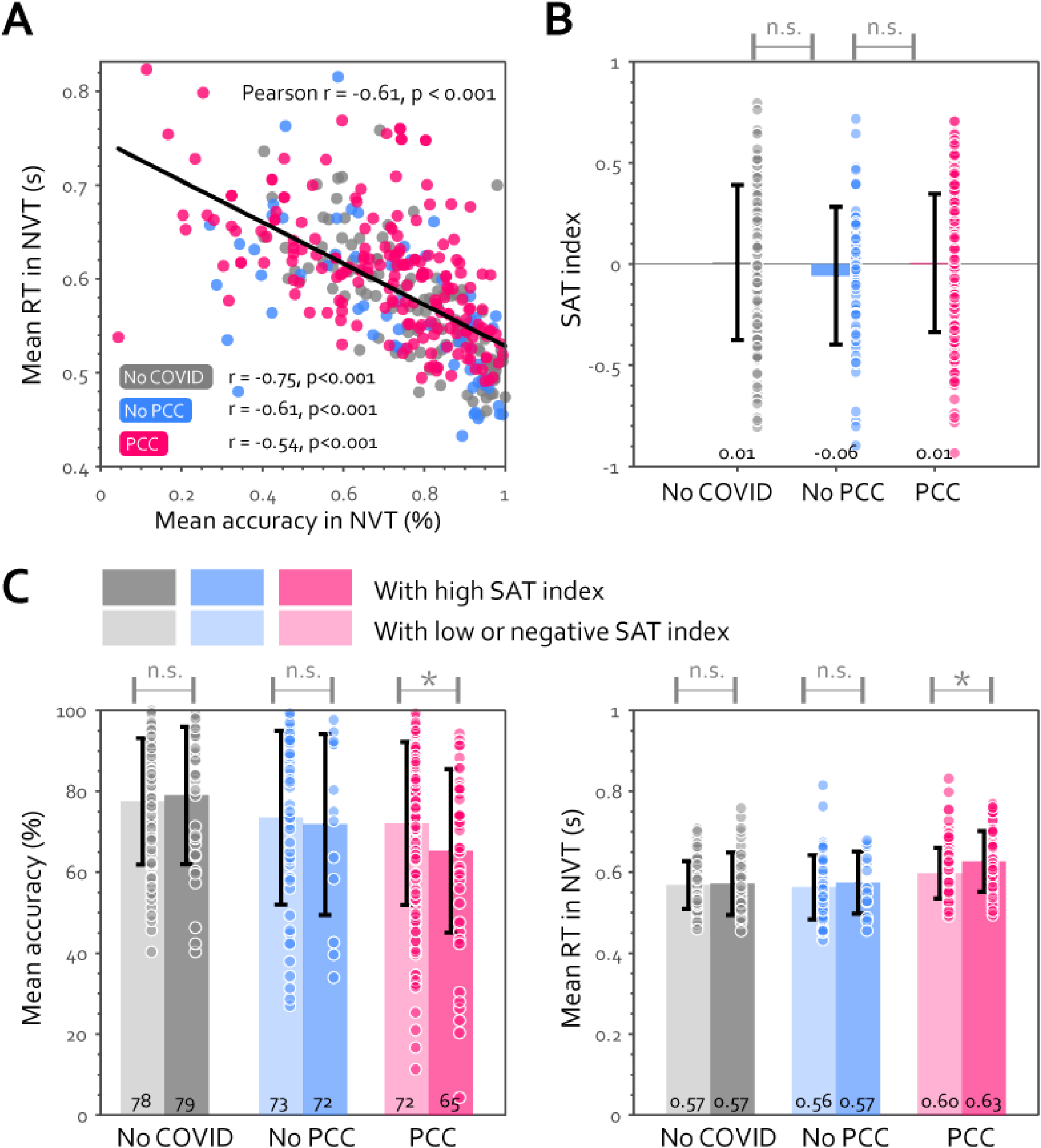
The slowness in PCC cannot be explained by speed-accuracy trade-off (SAT). (A) RT is negatively correlated with accuracy in the NVT across participants in every group. Pearson’s r and p values for all participants are shown at the top right and those for each group are shown at the bottom left of the graph. The SAT index for each group is shown in (B) with each dot representing individual data and the error bar showing ± 1 SD. SAT index for every participant is computed as the correlation coefficient of RT and accuracy for every minute. On the group level, PCC didn’t show any difference in the tendency to employ SAT in this test, as the SAT index is not significant above zero. (C) Comparing the objective performance within each group, between participants with high SAT index (i.e. median split of SAT index, shown in the right hand side dark colour bars) and the rest. n.s. means no significant difference. * means p<0.05.

One possibility for the good vigilance in PCC is that in order to reach good performance, patients worked harder. To examine this, we asked participants to rate their fatigue after every minute in NVT (“on-task tiredness”). PCC patients with normal response speed felt substantially more tired (55.3±2.2%) than other participants with normal speed (46.4±2.2%, t(265)=2.9, p=0.004, BF=6.4), suggesting they found sustaining attention on task more demanding.

### Cognitive slowing and depression distinguish PCC from healthy infected individuals

We then analysed which factors—mental health symptoms and objective cognitive metrics— were most effective in distinguishing infected individuals with PCC from other infected individuals without PCC, i.e., sensitivity for correctly assigning group membership to PCC. We selected all the metrics that showed significant group differences in this study and ranked them according to their importance in predicting PCC in infected individuals (i.e., PCC or No-PCC, **Figure 5A**). The rank represents the negative log of the p-values. Depression level was the best predictor, followed by RT in the SRT and in NVT. These three were significant predictors of the group (all p<0.0001), while PSQI and other key metrics in the NVT were not.

**Figure 5.**
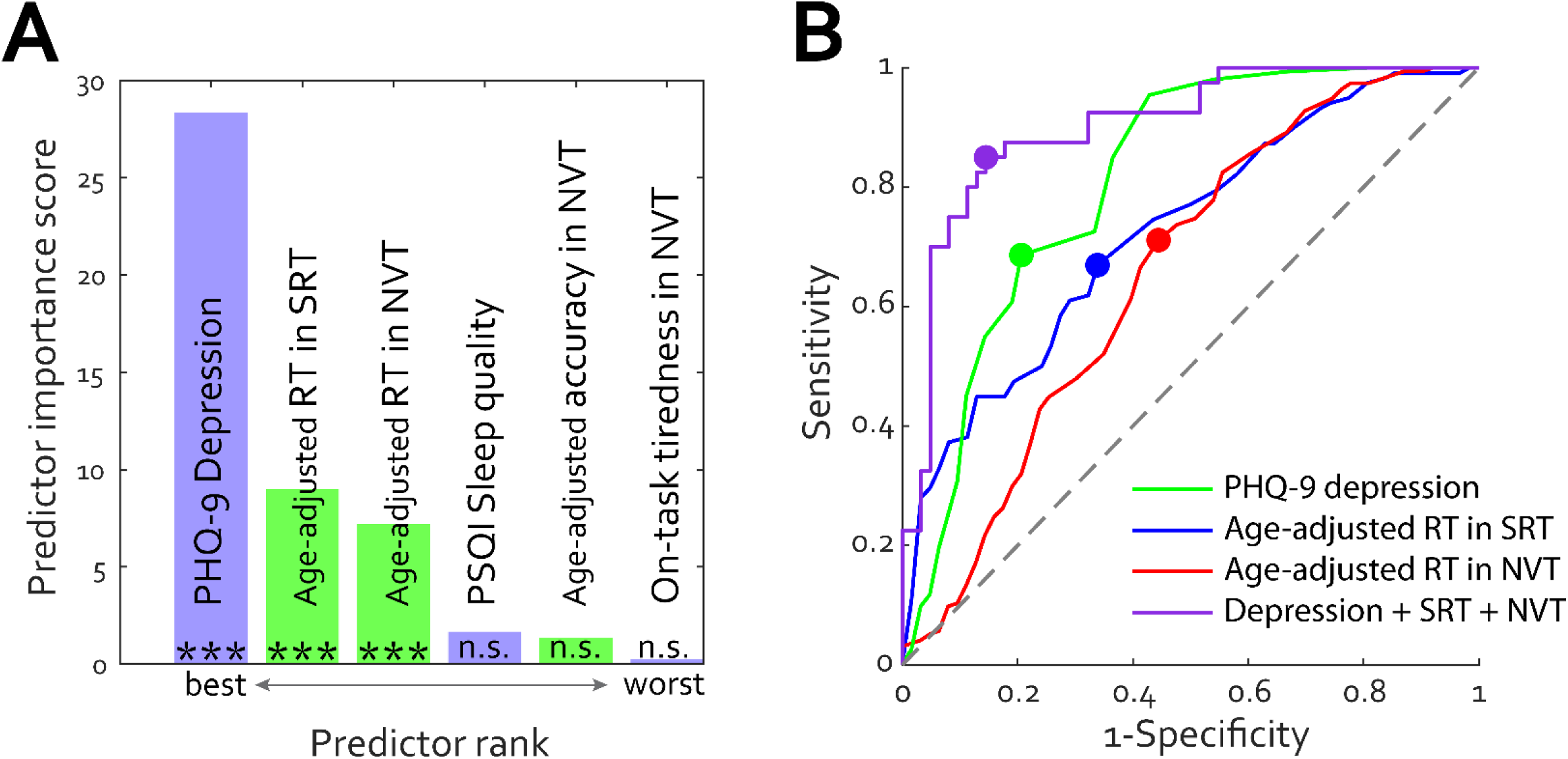
(A) Ranked measures in predicting group PCC or No-PCC. Cognitive measures are marked in lime, and self-reported measures (depression, sleep quality and on-task tiredness). *** indicates p-values below 0.001. n.s. means not significant. (B) Receiver Operating Characteristics (ROC) curves for group classification. The filled dots indicate the optimal operating points for each model. The purple curve represents the performance of a multiple logistic regression model with all three predictors, PHQ-9 derived depression level, age-adjusted RTs in SRT and NVT.

Figure 5B depicts the performance of these three significant predictors as ROC curves. PHQ-9 deprived depression scale (green curve) had a strong Area Under Curve (AUC) of 0.81, with 68.6% sensitivity and 79.4% specificity, at a cut-off score of 7 (i.e., if individuals scored PHQ-9 above 7). SRT (age-adjusted RT) could fairly distinguish individuals with PCC from those without PCC with 0.72 AUC, 66.9% sensitivity, and 66.1% specificity at the cut-off of 0.60 z (i.e., if individuals responded 0.6 SD slower than norm in SRT). NVT alone had slightly lower predicting performance (0.66 AUC, 71.1% sensitivity, and 55.6% specificity). We then used a multiple logistic regression with all three significant variables to classify PCC or No-PCC, which yielded an excellent AUC (0.90) and good sensitivity (85.0%) and specificity (85.5%) (purple curve in Figure 5B).

### Is cognitive slowing related to severity and time elapsed after acute COVID-19?

Figure 6A demonstrates that PCC patients hospitalised due to COVID-19 showed significantly lower accuracy in the NVT but no difference in RT in either task. Even after removing the cases with ICU admission (n=4), the WHO severity scale remained significantly negatively correlated with the mean accuracy in NVT (Kendall r=-0.18, p=0.007).

**Figure 6.**
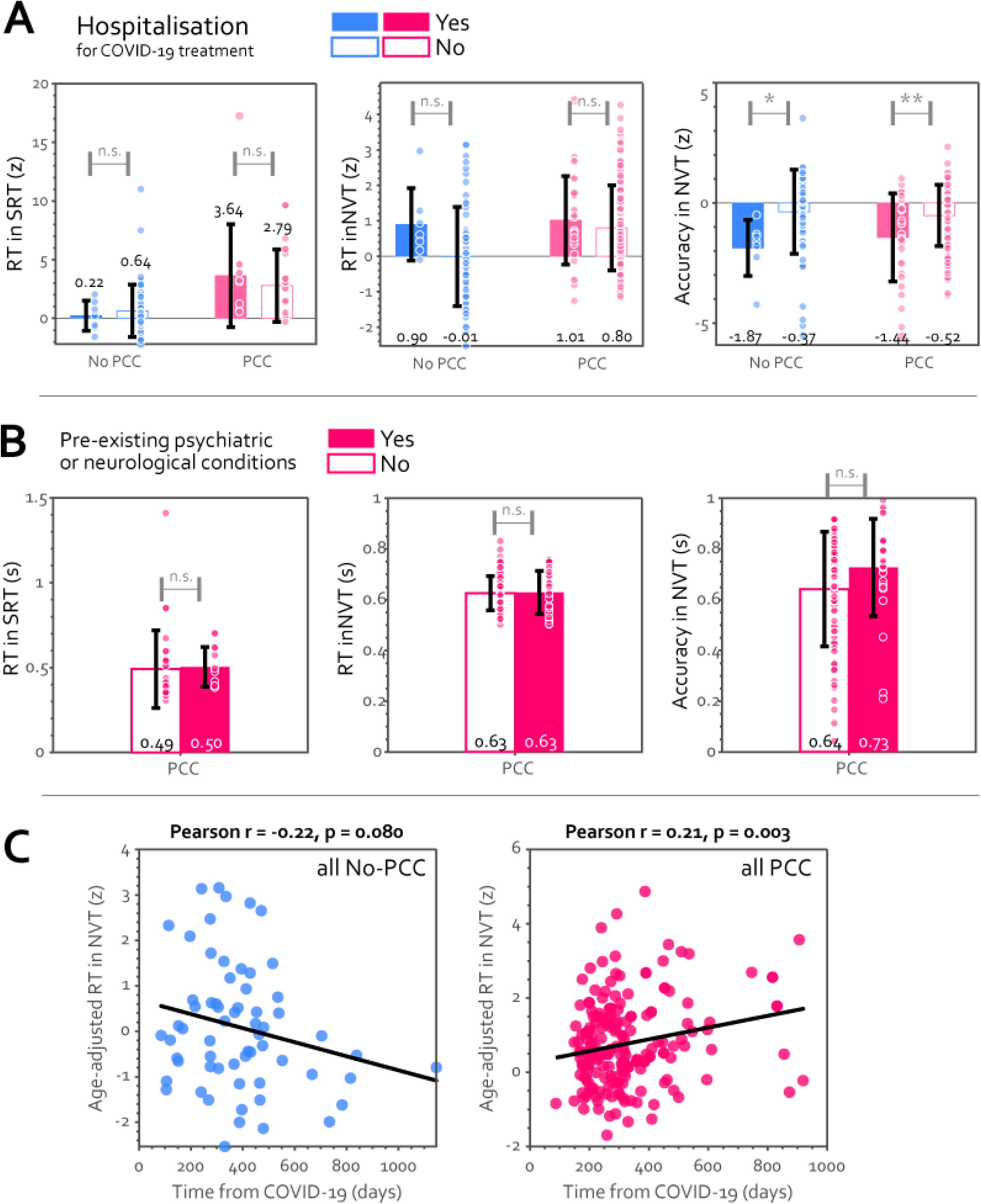
How did the acute COVID-19 infection affect objective performance? (A) Although hospitalised individuals, regardless of PCC status, demonstrated no difference in RT in SRT (left) or NVT (middle), they were significantly less accurate (right). (B) No difference was found between PCC patients with and without pre-existing psychiatric conditions prior to COVID-19. (C) In NVT, age-adjusted RT was marginally negatively correlated with time from infection in No-PCC participants, implying a gradual recovery (left). Cognitive slowing, however, was strongly positively correlated with time from infection in PCC group. n.s. means no significant difference. * means p<0.05, and ** means p<0.01.

Pre-existing psychological or neurological conditions did not differentiate PCC patients on objective performance neither (Figure 6B). Given that depression was the most prevalent of the pre-existing conditions here (45%, see Supplementary Materials), this result is consistent with the absence of the aforementioned relationship between depression and cognitive slowing in PCC.

Does the cognitive impairment get better with time? PCC patients showed the reverse trend: prolonged duration of PCC was linked with more severe cognitive slowing (r=0.21, p=0.003, Figure 6C).

## Discussion

The present study reported a significant psychomotor slowing in individuals diagnosed with PCC. Importantly, this cannot be attributed to poor global cognition as measured by a cognitive screening test (MoCA), fatigue, mental health-related symptoms, or speed-accuracy tradeoff. Additionally, the data indicate that this impairment does not improve over time. We also replicated this finding within each individual participant as well as with a separate cohort of PCC patients diagnosed by a different clinic located in a different country.

The existing body of research on chronic deficits in response speed exhibits significant inconsistencies. One of the earliest studies on post-COVID cognitive deficit suggested a substantial impairment in response speed in acute or sub-acute stage of COVID-19,^6^ while one investigation reported a modest deficiency only in severe or cognitive cases,^12^ and another online study reported no deficit in response speed in infected individuals with cognitive symptoms.^30^ In addition, Martin et al. found a deficit in perceptual processings speed in PCC patients with cognitive complaints.^10^

This finding offers an elucidation of the discrepancy by conducting a comparative analysis, for the first time, of the objective performance of individuals infected with and without PCC in relation to uninfected controls. Even though most (71.0%) of No PCC participants had normal SRT, this group had a higher prevalence (19.4%) of cognitive slowing compared with people without infection (4.0%, Figure 1E). The prevalence of severe cognitive slowing was even higher for PCC individuals (53%). In accordance with the fact that nearly two-thirds of the studies published up until February 2023 failed to adhere to any recognised deficits of PCC as outlined by authorities such as the NICE, CDC, and WHO,^14^ our finding suggests that the inconsistent findings concerning the persistent psychomotor deficit in post-COVID populations are likely due to heterogeneity in defining PCC in the published studies.

Depression and sleep deprivation may increase reaction time.^26–28^ However, our findings indicate that mental health symptoms alone cannot fully account for the cognitive slowing in PCC patients. This is in line with previous studies that found no correlation between the severity of mental health symptoms and chronic post-COVID cognitive deficit.^15,31–33^ Although depression on its own in PCC cannot explain cognitive slowing, Does this cognitive slowing resolve over time? Accumulating evidence suggests that the majority of individuals recover gradually after a mild-to-moderate COVID-19.^4^ However, the estimation of the duration of recovery is controversial, ranging from recovering within four months to not recovering two years after infection.^4^ Here we found that the cognitive slowing in PCC does not seem to resolve on its own. Instead of a gradual recovery (a negative relationship between time from infection and RT) an opposite trend was present amongst patients with PCC; patients who experienced PCC longer had more severe cognitive slowing. However, we must be cautious about the interpretation of the relationship with time since infection in the cross-sectional data. Specific variants of SARS-CoV-2 may be an important risk factor on cognitive slowing, as self-reported PCC symptoms are more common in the earlier waves before the Omicron variant.^3,34^ Thus, longitudinal studies with computerised speed tests in both PCC patients and those without PCC are needed to further confirm the group difference in relationship with time since infection.

Understanding of the underlying mechanisms responsible for the chronic cognitive deficit in PCC is still in its infancy, partly due to the lack of an objective signature in PCC. Here we identify a common cognitive deficit in PCC that can be quantitatively measured with an online platform, with all of our code publicly accessible.

## Contributors

SZ, KF, and MH conceived and planned the experiments. EMM, PAR, ASr, IU, VK, MR, SB, ASt, MS, EF and KF collected patient data. SZ, EMM, AS, AG collected control data. SZ, EMM and PAR contributed to data management and have directly access and verified the data. Statistical analyses were done by SZ, with input from EMM, KF, and MH. SZ, EMM, EF, KF and MH contributed to the interpretation of the results. SZ and MH wrote the manuscript with input from all authors.

## Data availability

De-identified data supporting this study may be shared based on reasonable written requests to the corresponding author. Access to de-identified data will require a Data Access Agreement and IRB clearance, which will be considered by the institutions who provided the data for this research.

The simple reaction time task and the number vigilance task can be tried online at [https://octalportal.com/pcc]. The source code is shared using a Creative Commons NC-ND 4.0 international licence upon reasonable written request to the corresponding author and publicly available at [https://octalportal.com/pcc].

## Acknowledgement

We would like to thank Ms Leanne Pridmore from Churchill Hospital in Oxford for helping us on data collection and Dr Sanjay Manohar for advice on statistical analysis.

This research was supported by funding from the Wellcome Trust, NIHR Oxford Health Biomedical Research Centre, and the Thüringer Aufbaubank (2021 FGI 0060). S.Z. and M.H. were funded by the Wellcome Trust (206330/Z/17/Z). E.M.M. was funded by Ph.D. scholarship “Landesgraduiertenstipendium” of Friedrich-Schiller-University Jena. K.F. was funded by German Forschungsgemeinschaft (DFG, FI 1424/2-1) and the Horizon 2020 Framework Programme of the European Union (ITN SmartAge, H2020-MSCA-ITN-2019-859890).

---

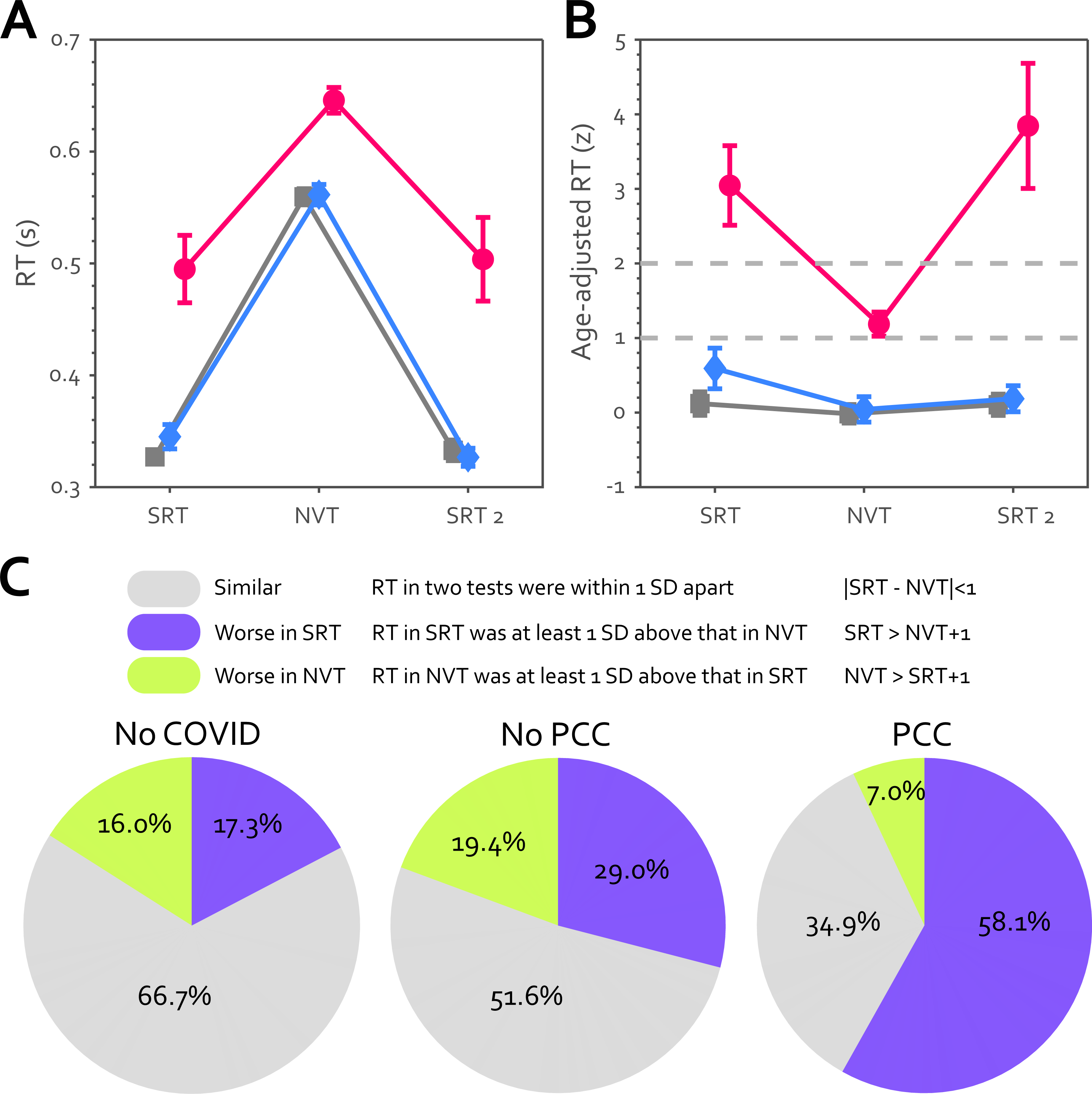

