## Supplementary Materials for "Long COVID is associated with severe cognitive slowing"

### Participants and demographic characteristics

245 PCC patients consented to participate in the study. After confirming the SARS-CoV-2 infection through a polymerase chain reaction test, patients sought treatment at the post-COVID outpatient clinic within the Department of Internal Medicine and the Neuro-Post-COVID-Centre within the Department of Neurology at Jena University Hospital, at least 12 weeks after the initial diagnosis. All patients fulfilled the NICE criteria for post-COVID syndrome.^1^

#### Patient exclusion criteria.

There were two exclusion criteria for PCC patients: (1) anyone who was (re-)infected with COVID-19 within 12 weeks of the day of testing. This is to avoid confounding the cognitive impairment during the acute phase of COVID-19. ^2–4^ 14 patients who alleged a recent re-infection within 12 weeks of the testing day were excluded. (2) anyone who failed to complete the NVT. 37 PCC patients (16.0% of 231=245-14) abandoned the Number Vigilance test prior to completion, with the majority leaving during practise. Compared with those who completed the test, these patients did not differ in gender (28 females (75.7%), χ2(1,N=171) = 0.062, p = 0.80), but, on average, they were older (age 55.4 (SD 12.0), range from 28 to 80, 75.7% were above 50; t(92) = 3.12, p = 0.002, BF10 = 15.36), had COVID-19 more recently (number of days since infection=256.3 (SD 152.3), range from 122 to 873, t(229)=-2.50, p=0.013, BF10=3.16) and had more severe acute symptoms (WHO severity scale: mean 2.91 (SD 1.51), range 1~6, t(96)=-2.46,p=0.016, BF=3.19) and also scored significantly lower on MoCA (26.5 (1.8), range 24~30, t(92)=-2.36, p=0.021, BF10=2.34). However, we do not believe this rate to be representative of the population as a whole. Although the tasks were self-administered, the researchers gave a slightly different instruction ("try this task, but feel free to quit if you find it difficult") at the start of the data collection phase, which unintentionally encouraged dropout. Then, we standardised the instructions given to participants during the recruitment phase. We now emphasise that the task is very challenging and encourage the participant to try to complete it. There were no more dropouts after the change of the pre-test instruction. These incomplete datasets were then excluded from the further analysis. After exclusion, there were 194 PCC patients remaining. Their demographics are shown in Table 1 in the main manuscript.

#### PCC Oxford group (PCC Ox)

In addition to the PCC group recruited in Jena, we also tested 77 PCC patients at Long COVID clinic in Oxford, UK to replicate the generalised slowness in SRT (see more details in Results). We did not have access to their patient information at the time this manuscript was written. One patient was excluded from analysis because we did not know their age or gender. The remaining 76 PCC patients had a mean age of 44.7 years (SD=12.7) and 52 (68.4%) were female. There was no difference in gender between the Jena and Oxford PCC groups (χ2(1,N=196)=0.9, p=0.34), but PCC Oxford patients were significantly younger (t(268)=2.85, p=0.005, BF=6.41). Considering the age effect on RT, it was not unexpected to find that the PCC Oxford group had slightly faster raw RT, but after accounting for the age effect (normalisation), there was no difference between these two groups (see Results).

#### Healthy controls

200 healthy participants were recruited in Jena (Germany), Oxford (United Kingdom), and via an online participant recruitment platform ([www.prolific.co](http://www.prolific.co)). The primary inclusion criteria for healthy participants were (self-reported) good health and absence of any neurological/psychiatric illness or other health concerns that could affect their cognition. All control participants were unaware of the purpose of this study. Immediately after completing the online tasks, participants were asked if they experienced brain fog or had any issue with concentration, then asked if they had contracted COVID-19 before and whether they were currently experiencing PCC.

13 participants who answered yes to the question “Do you consider yourself to have long COVID? (i.e., having COVID-related symptoms -- such as fatigue, brain fog -- more than 12 weeks)”, were classified as self-reported PCC. However, it was difficult to confirm whether these 13 participants met NICE criteria for post-COVID syndrome, like the PCC patients with an official diagnosis. Therefore, all self-reported PCC participants were excluded from further analysis. Only one participant withdrew out of the test and their information was also omitted from further analysis. Additional 11 participants were excluded due to recent COVID-19 infection.

After exclusion, we were left with 176 controls, who were then divided into two groups based on their previous COVID-19 infection status (yes or no to “Have you ever been infected COVID-19?”). No-COVID group included 113 participants who self-reported to be uninfected. No-PCC group included 63 participants who had COVID-19 more than 12 weeks ago but did not develop PCC. These two control groups did not differ in age (t(174)=0.26, p=0.80, BF10=0.18) or gender (𝜒2(1,N=88) = 2.0, p=0.16).

#### Demographics and medical history

For every participant, we know their age, gender, education level, time since COVID-19 and whether or not they experienced any concentration difficulty as part of PCC if applicable. There was no difference in age or gender. But PCC patients had fewer years in education than No-PCC or No-COVID groups (see Table 1). As expected, the prevalence of subjective concentration difficulty was significantly higher in PCC (69.6%) than No-PCC (23.8%) (χ2(1,N=150) = 41.0, p < 0.001).

We also acquired their hospitalisation history due to COVID-19 and WHO COVID-19 Severity Scale for all No-PCC participants and the vast majority of PCC patients (157 out of 194). These two groups had similar rates in hospitalisation (χ2(1,N=39) = 2.6, p=0.10) but PCC patients scored higher on WHO Severity Scale (t(154)=4.22, p < 0.001, BF10=458).

#### Pre-existing conditions

For 111 of 194 PCC patients, we also obtained their history of pre-existing neurological/psychiatric conditions. 31 of them (27.9%) had pre-existing neurological or psychiatric conditions. 16 of them had a history of depression (n=14) and/or anxiety (n=3). One had a suspected addiction to alcohol. One had a history of bulimia. Nine of them had various neurological conditions, for example, migraine, tinnitus, trigeminal neuralgia, rotatory vertigo, amyloid angiopathy, fibromyalgia, polyneuropathy. Unfortunately, we did not acquire the information about pre-existing conditions from our controls but based on their self-reported no history of neurological and psychiatric diseases, suggesting that they would have no pre-existing conditions.

### Experimental design

#### Simple Reaction Time Task (SRT)

A large red circle was present at the centre of the grey screen. The diameter of the circle was scaled as 50% of the screen height. Once participants pressed the spacebar, the red circle disappeared and would reappear after a randomised time interval between 0.5 and 2 seconds. There was a total of 16 trials, with the results of the first two trials omitted from further analysis.

This task was offered to a subset of participants, including 119 PCC patients (age 46.6 (SD 12.2, range 19 to 75, 80 females (67.2%)), 63 No-PCC participants (age 48.7 (10.5), 36 females), and 75 No-COVID participants (age 46.6 (11.9), 29 females).

#### Number Vigilance Test (NVT)

All participants completed the NVT, an online visual sustained attention task adapted from ^5^ and described in the preceding study.^6^ A German version, translated by KF, was provided for German speakers. The demo is available at [https://octalportal.com/pcc/] in both English and German.

This test was designed to examine visual sustained attention and measure vigilance decrement across minutes. We previously found that even young healthy people showed rapid decrement in accuracy over a few minutes. ^6^ In our previous study, 53 young No-PCC participants (i.e., infected with mild COVID-19 but did not experience post-COVID conditions) showed slightly but significantly worse accuracy at the end of the vigilance test compared to uninfected age-matched healthy controls.

In the original version of the task, a single digit (0-9) was presented at the centre of the screen for 50 milliseconds every second. Participants were instructed to press the spacebar on their keyboard as soon as they saw “0” (the target, presented randomly with a probability of 25%); no response was required for other digits. A semi-transparent jittered checkerboard pattern masked the digits.

During pilot testing, we found that the original task used for online young volunteers ^6^ might lead to frustration in clinical PCC patients due to its difficulty. Another recent work found that PCC patients (n = 40) needed more time to process multiple letters than uninfected controls, suggesting a general slowness in their visual processing. ^7^ Thus, here we extended the presentation duration of each digit from 50 to 100 milliseconds. Despite this adjustment, the current version of NVT was still demanding as even the uninfected group performed below ceiling in the first minutes on average (mean baseline accuracy=80.7±1.4%, t-test below 100% t(112)=-14.1,p<0.0001, BF10>1000) and 63.7% of uninfected participants performed below 90%.

The practice phase consisted of 90 trials, equivalent to 90 seconds and feedback (a tick or a cross) was provided after each trial. The first 12 practice trials were highly visible stimuli (not masked with semi-transparent checkerboards), and participants were required to get 100% accuracy to proceed. Subsequent non-practice trials and blocks contained no feedback. In total, each participant completed 540 trials, divided into 9 blocks, each containing 60 trials and lasting one minute.

#### Motivation and fatigue ratings with time on the NVT.

After each minute during testing, participants were asked to report their level of fatigue (“How tired do you feel now?”) and motivation (“How motivated do you feel?”) using a visual analogue scale. Responses were registered by clicking on the appropriate position on each scale. After completing all ratings, a “confirm” button appeared at the bottom of the screen, allowing participants to validate their ratings and start the next block. To control the time between blocks and to reduce variance between participants, a 15-second countdown timer was displayed at the top of the screen, and the next block would begin automatically once the timer lapsed.

#### Online testing

Both tasks were implemented using PsychoPy v2021.1.2. and hosted on the web-based platform Pavlovia (pavlovia.org). The choice of PsychoPy/PsychoJs was driven by its high timing precision for browser-based experiments. ^8^ We instructed all participants to use the Chrome browser on a desktop/laptop computer with a keyboard in order to minimise the potential differences in psychomotor response caused by way to response (keyboard, mouse clicking or finger tap) and devices (phone, iPad or laptop). However, the tasks can be completed on other devices and reaction times have been shown to be reliable across browsers. ^6,8^

#### Questionnaires

All participants completed two questionnaires for measuring depression level (Patient Health Questionnaire-9, PHQ-9) and their sleep quality (Pittsburgh Sleep Quality Index, PSQI).

For all PCC patients from Jena, the following mental health symptoms were also quantitatively measured using questionnaires:

- Anxiety was assessed by the anxiety subscale of the German version of the Hospital Anxiety and Depression Scale (HADS-D)^9,10^ which consists of 7 items. Scores range between 0 and 24 with higher scores indicating higher levels of anxiety.
- Depression was also assessed by the depression subscale of the German HADS^9,10^ that consists of 7 items. Scores range between 0 and 24 with higher scores indicating higher levels of depressive symptoms.
- Fatigue was assessed by two questionnaires: the Fatigue Assessment Scale (FAS)^11^ and the Brief Fatigue Inventory (BFI)^12^ with higher scores associated with higher levels of fatigue.
- Daytime sleepiness was measured by the German version of the Epworth Sleepiness Scale (ESS)^13^ consisting of 8 items asking for the probability of falling asleep from 1 (unlikely) to 3 (very likely) in 8 different situations. Scores range between 0 and 24 with higher scores indicating higher sleepiness and a score of ≥ 10 indicating excessive daytime sleepiness.
- Post-Traumatic Stress (PTSD) was assessed with the German version of the post-traumatic stress-scale-14 (PTSS-14).^14,15^
- Premorbid IQ was assessed using the Mehrfachwahl-Wortschatz-Intelligenztest (MWT-B).^16^ In this multiple-choice vocabulary test, participants recognised a German word from a list of four fictitious words. The sum of correctly recognised words (full score 37) was used as a measure of crystallised intelligence.
